# Exploring the Efficacy and Feasibility of using Biomarkers for Early Screening of Lung Cancer and their Potential Reverberation on Mortality Rate: A Review

**DOI:** 10.1101/2022.12.30.22284074

**Authors:** T Safari Vejin, J OLeary, L Singh, T Sullivan, S Aziague, A Akbariansaravi

**Affiliations:** American University of Antigua College of Medicine, Coolidge, Antigua and Barbuda New York Medical College School of Medicine, Valhalla, New York

**Keywords:** Lung cancer, Biomarkers, Standardizations, Screening methods, Prognosis

## Abstract

**Introduction:** Lung cancer patients often have a poor prognosis, due to various factors that play into the diagnosis, such as timeframe, early detection, and progression. The rate of patients with advanced-stage lung cancer is often high due to the inability to be diagnosed early. The lack of screening, therefore, increases the mortality rate of lung cancer. High mortality rate is also attributed to advanced lung cancer cases that are undiagnosed due to the lack of symptoms in patients early in disease acquisition.

**Methods:** This review explores lung cancer screening methods adopted around the world and those with the most promising standardizations. Criteria used to filter articles included lung cancer methods using biomarkers, research conducted in the United States within the past 10 years, screening methods observed in a clinical setting, and patients with primary lung cancer. Some practices that can be utilized to improve screening for lung cancer may involve using genetic markers to identify patients possessing genetic makeup associated with lung cancer.

**Results:** Our study included 32 articles in our review calculating the sensitivity and specificity for a variety of potential biomarkers including: 1) Volatile organic compounds has a specificity of 80% and sensitivity of 80% 2) Circulating tumor DNA has a sensitivity of 80% and specificity of 96% 3) miRNA molecules (miR-146-5p, miR-324-5p, miR-223-3p, and miR-223-5p) displayed sensitivities of 93%, 94%, 96%, and 95% respectively, with specificities of 33%, 23%, 27%, and 26% respectively 4) methylation beads specific genomic sequences (SOX17, TAC1, HOXA7, CDO1, HOXA9, and ZFP42) with sensitivities of 84%, 86%, 63%, 78%, 93%, and 87% respectively when found in sputum and 73%, 76%, 34%, 65%, 86%, and 84% respectively when found in serum. Specificities were calculated at 88%, 75%, 92%, 67%, 8%, and 63% respectively when found in sputum, and 84%, 78%, 92%, 74%, 46%, and 54% respectively in serum 5) Ga-Alfatide II scans were found to have a sensitivity of 86% and a specificity of 85% 6) Combined score had a sensitivity of 78% and specificity of 50% 7) WT-IDH had a sensitivity of 63% and specificity of 93%.

**Discussion:** In such a high-risk patient population, cautious preventative measures can be used such as lifestyle choices and environmental exposures that are known risk factors. Informed physicians will be able to provide care directed toward minimizing patient’s modifiable risk factors to lower incidence rate and routine imaging offered when there is an index of suspicion to minimize mortality rate in high-risk populations. This review highlighted practices such as circulating tumor DNA and a Ga-Alfatide II CT/PET scan that can be utilized to improve screening in the future and further dissected errors made in lung cancer diagnosis.

## Introduction

There have been many advances regarding the diagnosis and treatment of lung cancer. Despite these changes, the progress in the field of primary respiratory neoplasm remains incomparable to the constant improvements observed in other oncological fields, such as OB/GYN and Gastroenterology. Considering the lack of advancements, the number of patients affected by lung cancer remains unchanged and has been so for the past decade, suggesting little improvement in medical developments for lung cancer. Since 1987, lung cancer remains the leading cause of death in both men and women. The alarming rate of lung cancer deaths has also prompted findings exploring methods that could be used in lowering lung cancer death rates, as well as disease prevention. The death toll for lung cancer peaked in 2005 and decreased by a mere 10,000 by 2016, further emphasizing the minimal rate of improvement over a span of 10 years.^1^ The continued incline of disease progression coupled with the lack of improvement has caused a rise in lung cancer mortality. In 2018, the incidence of lung cancer worldwide was 2.1 million new cases per year.^2^ In addition, it was calculated that 1.8 million people will succumb to their lung cancer diagnosis that same year.^3^ The vital question posed in the data addresses the comparable disparity between the high mortality rate and low incidence rate of lung cancer.

Improvement can be seen in cases of lung cancer by targeting three vital entities involved with chronic conditions—primary, secondary, and tertiary prevention. Primary prevention methods include behaviors involved in preventing the condition from developing, as well as environmental and genetic contributing risk factors. Furthermore, primary prevention acts as the first shield in disease deterrence by exploring ways by which lung cancer incidence can be lowered. Failure of primary prevention leads to the adoption of secondary prevention, which aids in screening with the inclusion of symptom management. Tertiary prevention is utilized in advanced disease states and explores treatment methods, including their success rates. Of the three prevention methods, primary prevention remains understudied in data and literature. By employing these prevention techniques, early detection of lung cancer can be further improved and better studied. Affordability plays an important role in early detection, including clinical method standardization, as it could increase the probability of early cancer detection during general checkups with primary care physicians.

Disease burden is higher in the African American population compared to other races, indicating an immense racial disparity concerning lung cancer incidence. Lung cancer incidence is 30% higher in African American men compared to Caucasian men; although, the overall smoke exposure and risk factors of the former were lower.^4^ A similar pattern is also observed for women; however, incidence has been reported to be the same amongst African American women and Caucasian women, with lower exposure observed in African American women. ^5^ Regarding gender differences, men have been reported to have a higher incidence rate for lung cancer, whereas women are reported to have a higher prevalence.^5^ Incidence rates exploring age demographics have been lowered from 42-years-old to 36-years-old for diagnosis.^5^

High mortality rate can be mainly attributed to squamous small cell and adenocarcinoma.^3^ Small cell lung carcinoma (SCLC), also referred to as oat cell carcinoma makes up 20% of respiratory cancers.^3^ SCLC consists of a gray-white central mass containing small round blue (polygonal) cells in clusters. Being the most aggressive respiratory cancer due to its rapid growth and early dissemination, SCLC depicts a poor prognosis. In addition, SCLC cells are notorious for their neurosecretory granules that give rise to a myriad of paraneoplastic syndromes, including Syndrome of Inappropriate Antidiuretic Hormone (SIADH), Horner’s Syndrome, Lambert Eaton Syndrome, and Cushing’s Syndrome. Exceptions concerning paraneoplastic syndromes associated with SCLC include hypercalcemia and PTrH secretion, which are involved in Squamous Cell Carcinoma (SCC), as well as high HCG levels seen in LCCs. LCCs present with large anaplastic cells that are poorly differentiated. Furthermore, the secretion of HCG in LCCs leads to symptoms of gynecomastia in both men and women, in addition to a false positive pregnancy test in either gender.^4^

SCLC cells are of neurosecretory origin and function as neuroendocrine tumors. They also stain positive with Synaptophysin, Chromogranin, and Bombesin stains. SCLC is associated with the L-myc gene mutation, which is a basic helix-loop-helix transcription factor that is amplified and overexpressed in SCLCs.^3^ Current chemotherapy treatment of SCLCs involves the use of monoclonal antibodies which target the myc homolog sequence box and prevent its expression.^4^ The poor prognosis in these cancers, including recently discovered gene mutations observed in adenocarcinoma are being targeted by biomarker chemotherapy techniques.

Adenocarcinomas comprise 35% of all respiratory cancers.^3^ Contrary to other respiratory carcinomas, there has been a stronger correlation observed with non-smokers with a higher prevalence noted in women compared to men. Regarding frequency, non-smokers have a 25% incidence rate worldwide generally with all cancer types.^5^ Adenocarcinomas present as a gray-white peripheral mass organized in a lepidic pattern in the lungs. Furthermore, adenocarcinomas consist of mucous-secreting glands accompanied by pleural puckering with a tendency to form scarring. These types of scarring are also referred to as scar carcinomas. Primary adenocarcinomas have a better prognosis compared to their lung tumor counterparts, presenting with the best survival rate strongly dependent on early surgical resection of the tumor.

Specific gene mutations, such as P53 and retinoblastoma have been strongly associated with lung cancers overall.^1,2,4^ Adenocarcinomas have been associated with numerous gain-of-function gene mutations, involving tyrosine kinase receptors, including EGF, ALK, ROS, MET, and RET.^1,2,4^ These carcinomas were previously identified to involve the K-ras mutation; however, with more current studies, roughly 10% have been identified to have EGFR mutation and 5%, the EML4-ALK fusion protein mutation.^1,2,4^ ALK is a gene that codes for a receptor tyrosine kinase activated by the EML gene. Upon fusion of the ALK and EML genes, the receptor tyrosine kinase acquires uninhibited activity, which creates the basis for the formation of peripheral mass adenocarcinoma. Despite association with multiple gene mutations, the EGFR and EML4-ALK fusion mutations have been targeted by specific small-molecule inhibitors used as chemotherapy treatment.^1,2,4^

**Figure 1:**
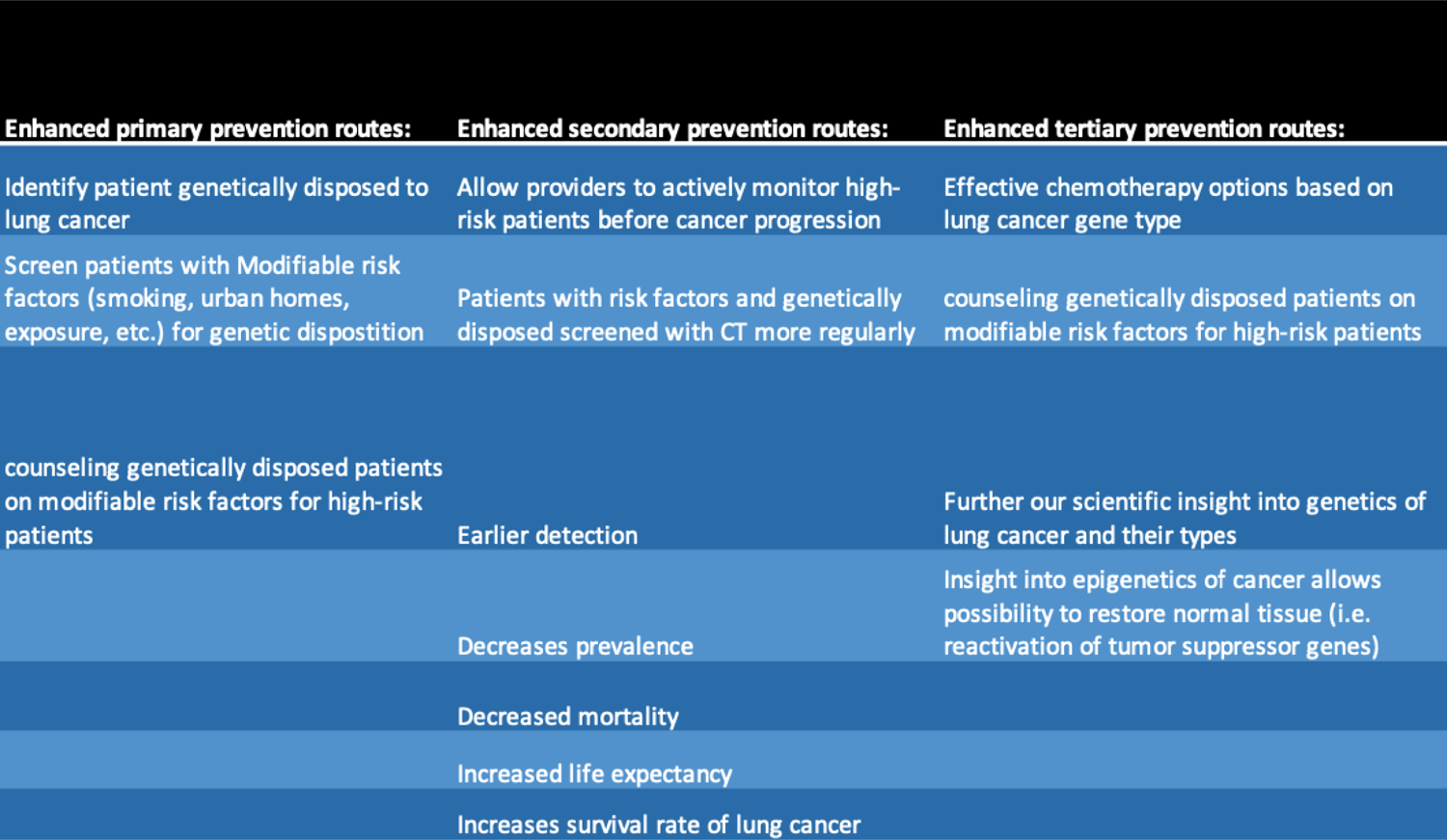
Table of utilities for epigenetics in lung cancer screening and treatment

Peripheral Non-small cell carcinomas (NSCCs) with EGFR mutations have shown promising results when treated with Erlotinib, Gefitinib, and Erlotinib.^3,4,5^ These chemotherapy treatments target inhibition of angiogenesis and epidermal growth factor activity in tumor cells, as well as inhibited tyrosine kinase activity. In addition, the adenocarcinomas that contain the EML4-ALK mutation have been targeted by Crizotinib as chemotherapy.^3,4,5^ Crizotinib is an anaplastic lymphoma kinase (ALK) and C-ros oncogene 1 (ROS1) inhibitor that targets inhibition of ALK phosphorylation and signal transduction of the NSCC tumor cells, inhibiting their growth at the G1-S cell cycle phase.^3,4,5^ Adenocarcinomas have shown a high preference for metastasis to the lung, with far more occurrence of metastatic adenocarcinomas involving the lung than primary adenocarcinomas.^3,4,5^

Squamous cell carcinoma and small cell carcinoma are strongly associated with smoking with a higher prevalence in males. Compared to adenocarcinoma or large cell carcinoma, which targets the periphery of the lungs, both squamous and small cell carcinomas are central masses. Squamous cell carcinoma accounts for 30% of all lung cancer incidents, with a higher prevalence in males and a strong correlation to smoking history.^1^ It originates from the bronchial epithelium after progressively evolving from metaplasia to dysplasia and ultimately becoming its invasive counterpart, carcinoma in situ.^1^

Bronchogenic carcinomas comprise 5% of all lung cancers and originate from terminal bronchioles.^1,2,4^ These carcinomas further develop as columnar tumor cells that proliferate along the alveolar walls, forming endobronchial lesions. They form gray-white mucinous nodules in the periphery of the lungs. Bronchogenic carcinomas have been identified as the main contributors to mortality amongst male smokers aged 50-80 and have rapidly become the main cause of death amongst female smokers in recent years.^1,2,4^ The risk factors associated with bronchogenic carcinomas include cigarette smoking, occupational exposure to toxins, such as asbestos & uranium mining, radiation, and air pollution. The presence of pre-existing chronic fibrotic lung disease further worsens the symptoms of bronchogenic carcinomas. Chronic fibrotic lung disease that has been associated with the onset of bronchogenic carcinomas include idiopathic pulmonary fibrosis, non-specific interstitial pneumonia, cryptogenic organizing pneumonia, various pneumoconiosis, and collagen vascular diseases such as Systemic Lupus Erythematosus (SLE) and Rheumatoid Arthritis (RA).^1,2,4^

The current golden standard for lung cancer screening involves the administration of low-dose lung CT to patients between the ages of 50-80.^7^ This screening method can be inaccurate due to risks associated, which include false positives & overdiagnosis, which could limit chances of early detection.^6^ The current protocol for the treatment of non-small cell carcinomas is surgery. On the other hand, chemotherapy is the best option in the case of unresectable tumors, such as small cell carcinomas. Despite chemotherapy treatment for patients with small cell carcinomas, the prognosis is extremely poor and little improvement has been shown in the past decade.^6^ That being said, roughly 10% of patients have a 5-year survival rate.^7^ There has been a plateau in the advancements of screening and treatment of lung cancer compared to other oncological areas. The main topic of importance in this paper highlights the importance of using biomarkers that are targeted for a specific gene mutation, which either predisposes or plays a role in the development of the different kinds of lung cancers.

## Methodology

### 1.1 Method

The initial search on PubMed using the keywords mentioned in the methodology yielded 32 articles. These articles were studies conducted over a span of 5 years. Due to the narrow timeframe, an additional search was performed to include articles published within the past 10 years. By broadening the search, 32 additional articles were discovered. A total of 61 articles published within the past 10 years on PubMed matched the criteria.

### 1.2 Criteria

The literature search on lung cancer screening involved the main scholarly platforms utilizing the same keywords across all databases. The keyword used across all platforms was “<u>biomarkers for early diagnosis of lung cancer</u>”. This allowed for the extraction of many articles (n =2,109), however we were not selective for any specific biomarkers or subtypes of lungs cancer. The articles extracted were then filtered concisely using the following criteria: 1) the article was centered around clinical research 2) the research was conducted within the last 10 years 3) the research was completed within the United States 4) the article compared the amount of research conducted in the United States versus other countries 5) the article included the exploration of future directions of the study 6) the article involved analysis of international research trials and reviews 7) the article permitted assessment of risk factors across countries and cultures. A final record of 49 research articles fit the final definitive criteria and were included in our review. The search was executed exclusively on PubMed.

This review encompassed study criteria involved in the analysis of biomarker screening methods for patients with a variety of cancers, including lung cancer. Applicable data for each study regarding lung cancer was carefully organized throughout the review. The table below gives a good summary of the data collected during the literature review.

### 2.3 Defining characteristics across articles

We evaluated successful biomarkers pertinent to lung cancer screening across the articles reviewed. To compare the efficacy of biomarkers used in the early detection of lung cancer, Positive Predictive Value (PPV) and Negative Predictive Value (NPV) were considered. Patients’ demographics in addition to risk factors pertinent to specific lung cancer types were further evaluated as part of representative data. The collection of the aforementioned data was correlated to various routes of lung cancer prevention with consideration to biomarker screening techniques. PPV, sensitivity, and specificity were taken as a collective average from all articles and represented in this study. Articles that did not contain these statistics were excluded during calculations of the average.

### 2.4 Future approach

Further analysis included identifying the lack of data and inconsistencies across the journals.^3^ In addition, data represented in Table 1 were visually displayed with highlights of significant findings. The visual included categories such as demographics, risk factors, lung cancer types, and biomarkers. The future direction of the paper was obtained based on data findings and possible improvements in the clinical implementation of biomarkers. Additionally, the lack of important data in lung cancer literature, involving screening methods are valuable considerations that should be analyzed in future studies and journals.

**Table 1:** List of standardized information that was collected for all the articles meeting the criteria

|  |
| --- |
| Stanford information of the article; Name and authors, year published |
| Early screening method used |
| Number of participants |
| Type of lung cancer studied (include breakdown of the lung cancer types; adenocarcinoma, squamous cell carcinoma and small cell carcinoma of the lungs) |
| Biomarkers used |
| Number of patients detected using the screening method |
| Breakdown of demographic distribution of patient population used in that study; ethnicity, age, state, rural vs. urban residence, exposure to known lung cancer carcinogens (radon, smoke, pollution, asbestos, prev. tuberculosis infections) |
| Associations with paraneoplastic syndromes and other possible early manifestations |
| Concluded statistics calculated for the study: sensitivity and specificity for group cancers, PPV |
| Was a biomarker shown to be more effective towards a certain type of lung cancer over others (included for studies including patients with all 3 main types of lung cancer) |
| Additional notes/notable findings |
| Future direction suggested in the paper |

## Results

### 1.3 Collection of identifying variables comparing various types of lung cancers

The following are observations summarized in Table 2 with calculations for sensitivity and specificity summarized in Table 3. Research published between 2011 and 2020 demonstrates the efficacy of biomarkers by comparing types of lung cancer, mainly adenocarcinoma, and squamous cell carcinoma. Data was collected highlighting screening methods and improvements in biomarkers with consideration to study participants’ age and smoking history. Screening methods mainly included chest x-ray, PET/CT scan, blood samples, sputum samples. Most of the studies between the years observed showed that biomarkers enhanced the sensitivity and specificity greatly in elderly patients that had a history of smoking.

**Table 2:**
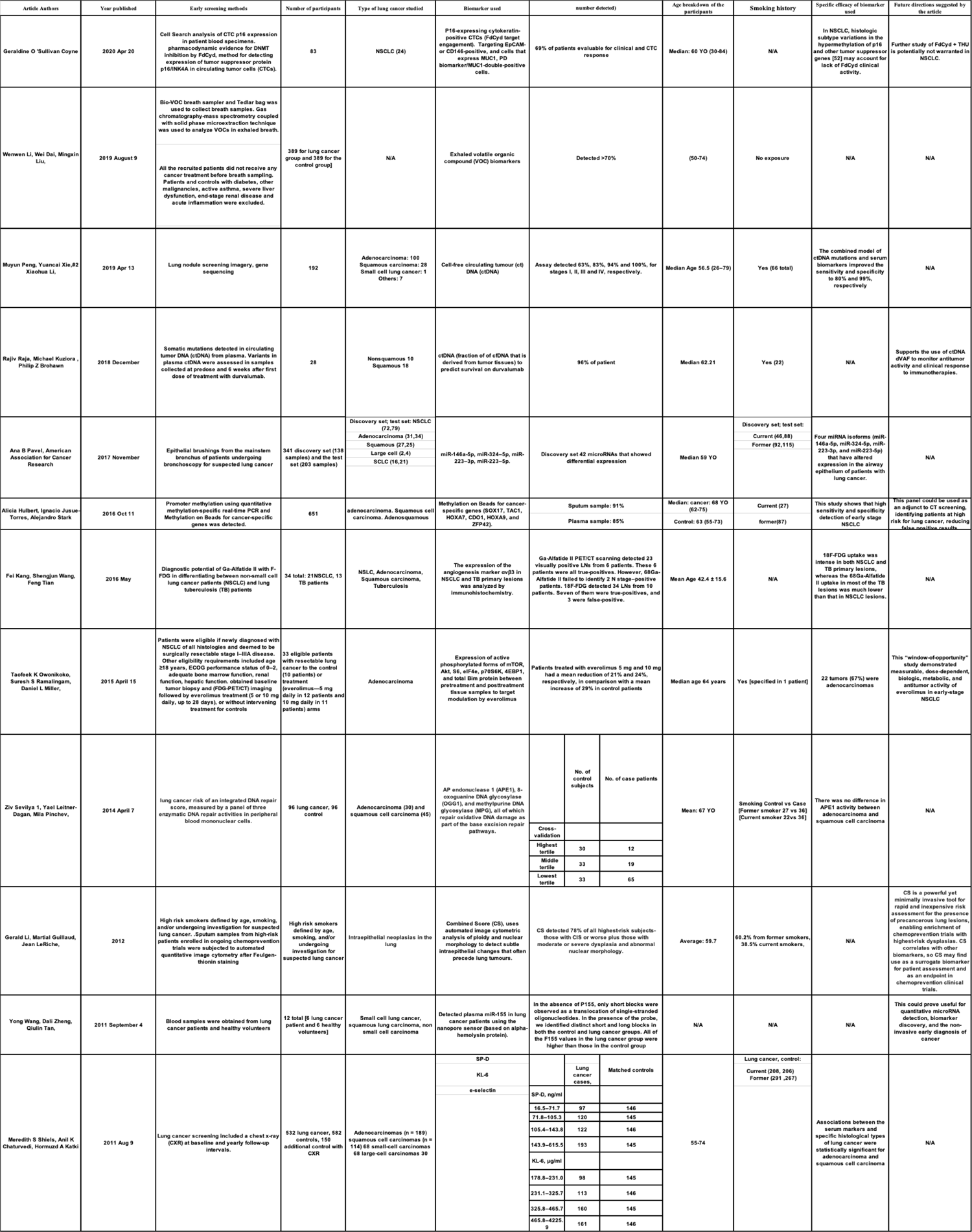
Data collected from each article according to SC vs. NSC lung cancer and its subtypes

**Table 3:** Table of useful biomarkers found in systematic review for potential for detection & therapy of lung cancer

| Biomarker | Sensitivity | Specificity |
| --- | --- | --- |
| Exhaled volatile organic compound (VOC) biomarkers | 80% | 80% |
| Cell-free circulating tumour (ct) DNA (ctDNA) | 80% | 96% |
| miR-146a-5p, miR-324-5p, miR-223-3p, miR-223-5p. | mRNA biomarker (0.94), mRNA biomarker + miR-146a-5p (0.93), mRNA biomarker + miR-324-5p (0.94), mRNA biomarker + miR-223-3p (0.96), mRNA biomarker + miR-223-5p (0.95) | mRNA biomarker (0.27), mRNA biomarker + miR-146a-5p (0.33), mRNA biomarker + miR-324-5p (0.23), mRNA biomarker + miR-223-3p (0.27), mRNA biomarker + miR-223-5p (0.26) |
| Methylation on Beads for cancer-specific genes (SOX17, TAC1, HOXA7, CDO1, HOXA9, and ZFP42). | Sensitivity in sputum: SOX17(84), TAC1(86), HOXA7(63), CDO1(78), HOXA9(93), ZFP42(87). Sensitivity in plasma: SOX17(73), TAC1(76), HOXA7(34), CDO1(65), HOXA9(86), ZFP42(84) | Specificity in sputum: SOX17(88), TAC1(75), HOXA7(92), CDO1(67), HOXA9(8), ZFP42(63). Specificity in plasma: SOX17(84), TAC1(78), HOXA7(92), CDO1(74), HOXA9 (46), ZFP42(54) |
| Ga-Alfatide II | 85.71% | 84.62% |
| Combined Score (CS) | 78% | 50% |

In addition, the cohort study calculations in the screening for isocitrate dehydrogenase (IDH) values are summarized in Table 4. The data demonstrates the efficacy of screening for IDH in serum by comparing a training cohort with a validation cohort using a total of 1166 participants in the diagnosis of non-small cell lung cancer. Finally, demographics based on gender and age are also displayed with the latter being summarized by median, interquartile range, and range of the study population.

**Tables 4a and 4b:** Utilization of isocitrate dehydrogenase 1 as a serum protein biomarker for the early detection of non-small-cell lung cancer: A multicenter in vitro diagnostic clinical trial^7^

|  | AUC (95% CI) | Se% | Sp% | PPV% | NPV% | Positive LR | Negative LR |
| --- | --- | --- | --- | --- | --- | --- | --- |
| Training cohort |  |  |  |  |  |  |  |
| NSCLC vs HC, BPC and OC | 0.870 (0.840-0.899) | 63.3 | 86.8 | 87.1 | 62.7 | 4.80 | 0.42 |
| NSCLC vs HC | 0.915 (0.887-0.942) | 63.3 | 92.9 | 95.4 | 52.0 | 8.91 | 0.40 |
| Stage 0-IA NSCLC vs HC, BPC and OC | 0.859 (0.826-0.892) | 58.6 | 86.8 | 79.3 | 70.9 | 4.44 | 0.48 |
| Stage 0-IA NSCLC vs HC | 0.907 (0.875-0.938) | 58.6 | 92.9 | 92.2 | 61.0 | 8.25 | 0.45 |
| Stage IB-IV NSCLC vs HC, BPC and OC | 0.886 (0.852-0.919) | 70.7 | 86.8 | 74.4 | 84.5 | 5.37 | 0.34 |
| Stage IB-IV NSCLC vs HC | 0.927 (0.896-0.958) | 70.7 | 92.9 | 90.0 | 77.8 | 9.96 | 0.32 |
| Validation cohort |  |  |  |  |  |  |  |
| NSCLC vs HC, BPC and OC | 0.745 (0.704-0.786) | 55.0 | 86.3 | 82.8 | 61.5 | 4.01 | 0.52 |
| NSCLC vs HC | 0.730 (0.684-0.776) | 55.0 | 89.3 | 92.7 | 44.6 | 5.12 | 0.50 |
| Stage 0-IA NSCLC vs HC, BPC and OC | 0.797 (0.730-0.865) | 59.1 | 86.3 | 53.4 | 88.8 | 4.31 | 0.47 |
| Stage 0-IA NSCLC vs HC | 0.788 (0.711-0.865) | 59.1 | 89.3 | 75.0 | 80.0 | 5.50 | 0.46 |
| Stage IB-IV NSCLC vs HC, BPC and OC | 0.746 (0.697-0.796) | 54.4 | 86.3 | 74.2 | 72.3 | 3.97 | 0.53 |
| Stage IB-IV NSCLC vs HC | 0.732 (0.676-0.788) | 54.4 | 89.3 | 88.3 | 56.8 | 5.07 | 0.51 |

AUC, area under the receiver operating characteristic curve; BPC, benign pulmonary condition; HC, healthy control; LR, likelihood ratio; NPV, negative predictive value; OC, other cancer; PPV, positive predictive value; Se, Sensitivity; Sp, Specificity.

|  | Training cohort (620 cases) |  |  | Validation cohort (546 cases) |  |  |
| --- | --- | --- | --- | --- | --- | --- |
|  | NSCLC patients | HCs | Disease controls (patients with BPCs or OCs) | NSCLC patients | HCs | Disease controls (patients with BPCs or OCs) |
| No. | 362 | 155 | 103 | 298 | 121 | 127 |
| Age, y |  |  |  |  |  |  |
| Median | 59.0 | 37.0 | 53.0 | 61.0 | 34.0 | 59.0 |
| (Q1, Q3) | (53, 65) | (31, 46) | (45, 64) | (54, 68) | (27, 45) | (50, 67) |
| Range | 35-82 | 23-67 | 26-84 | 26-85 | 18-72 | 17-86 |
| Sex |  |  |  |  |  |  |
| Female/ male | 201/161 | 87/68 | 49/54 | 120/178 | 64/57 | 47/80 |

## Discussion

Early detection remains one of the best ways to reduce lung cancer deaths. By detecting lung cancer in its early stages and resecting it, patients will be able to have a better prognosis. Breast and Prostate cancers are the most prevalent, but not the deadliest. Inversely, lung cancer is less common with the highest rates of death. Lung cancer remains the leading cause of cancer deaths, mainly due to underdeveloped screening mechanisms. This is attributed to the high association of smoking with lung cancer. It is important for medical caregivers to target smoking cessation in patients to further reduce the risk of developing lung cancer.

In contrast to lung cancer, there are various screening methods available for other cancer types. Screening allows for early detection, which reduces mortality rates. For example, breast cancer screening includes mammograms and physical examinations. Prostate cancer screening methods involve Prostate-Specific Antigen (PSA) and Digital Rectal Examination (DRE). Colorectal cancer screening includes hemoccult testing of the stool and colonoscopy. These cancers have higher incidences compared to lung cancer, but their screening techniques led to a lower mortality rate. Lung cancer survival rates remain at 15% when comparing data from roughly 30 years ago and data from the past 5 years. Developments in screening methods for lung cancer will improve survival rates and a variety have been analyzed during our investigation.

Volatile organic compounds (VOC) can be measured using exhaled breath analysis making the procedure completely non-invasive, giving it great potential as a screening method for lung cancer^23^. Our study found that currently VOC exhaled breath analysis has both a sensitivity and specificity of 80%, which is on the border of being a viable screening test. However, there are issues with using VOC as a potential biomarker. For instance, there are no unified VOC biomarkers for lung cancer^23^, thus further investigation must be conducted to determine the exact VOC biomarkers found in lung cancer. Such investigations will raise the sensitivity and specificity and could potentially make VOC a gold standard for lung cancer screening.

Combined score (CS) analysis refers to the correlation between several lung cancer risk factors including histopathological grade, age, smoking status, and p53 and Ki67 immunostaining^21^. CS is an incredibly powerful tool having the benefit of being minimally invasive and accounting for multiple biomarkers associated with lung cancer. However, our study found that CS has a sensitivity of 78% and specificity of 50%, showing that CS requires further investigation into the risk factors the test will analyze. With a specificity and sensitivity that low, CS is currently not a candidate as a screening method, however there is much potential. Should further investigation into CS alter the risk factors to more accurate and precise variables, the sensitivity and specificity would increase and its possible CS could become a viable screening technique miRNAs are a type of nucleic acid represented as small, non-coding RNA molecules binding to their complementary targets of 3’UTR segments in RNA; repressing gene expression and protein translation. However, unlike mRNA they are much more stable molecules, and thus much more easily measured in degraded tissue such as those caused by smoking^30^. Our study identified four miRNA isoforms (miR-146a, miR-324-5p, miR-223-3p, and miR-223-5p) which have altered expression in epithelial airways of lung cancer. As compared with normal tissue, all four miRNA molecules have down-regulated expression and in addition, were found to be implicated in tumor suppressive pathways^30^. Our study found that the down regulation is significant enough for the screening test for each of these four miRNAs to each have a sensitivity of 93% or higher, an indication of a valid screening test. However, each of the four miRNAs listed above has a specificity of 33% or lower, which could be attributed to their role in tumor suppression. While these molecules are excellent at detecting cancer, they are terrible at preventing false-positives, thus using miRNA molecules as a potential screening test is not possible at this time.

Isocitrate dehydrogenase I (IDH) is a catalytic enzyme found in the TCA cycle found in the cytoplasm and peroxisomes, catalyzing the oxidative decarboxylation of isocitrate to alpha-ketogluterate. IDH1 is an extremely important enzyme as it produces NADPH in the cytoplasm and has been getting widespread attention due to the Arg-132 mutation^7^. A mutation that causes the generation of the metabolite 2-hydroxygluterate leading to a decrease in the original product.

The metabolite will suppress activity of the enzyme prolyl hydroxylase and prevents degradation of hypoxia inducing factor resulting in 70% of grade II/III gliomas, 8.5% of acute myeloid leukemia, as well as a few other types of tumors^7^. While there has been no known association with lung cancer, prometric discovery assays identified that WT IDH1 is overexpressed in non-small cell lung cancer and that IDH1 can promote the proliferation of this cancer line^7^. However, our study found that the sensitivity is not above 80% and the specificity is not above 70%, thus WT IDH1 is a poor screening method due to the large proportion of false-negatives and false-positives

Genome methylation is a potential screening technique as the methyl group stabilizes the gene and thus potentially increases sensitivity^18^. In addition, our study found six genes (SOX17, TAC1 HOXA7, CD01, HOXA9, ZFP42) that were identified as having a high frequency of altered DNA methylation in lung squamous and adenocarcinoma, but not in normal lung tissue^18^. While a promising method of lung cancer screening, as the sample collection is from either serum or sputum and thus non-invasive, there are several issues that must be addressed. There are some patients with undetectable DNA methylation in either collection sample (however sputum samples were slightly higher)^18^. Unfortunately, our study found this effect to have an effect on sensitivity and specificity as neither of these genes had both of these values above 80%. While this technique remains promising, further investigation into DNA methylation in lung cancer will need to be conducted for detection of methylation beads on genes to be a viable lung cancer screening.

Circulating tumor DNA (ctDNA) is extremely specific for a patient’s tumor due to somatic mutations in tumor DNA that can be identified from normal DNA^32^. The benefit of ctDNA is the reduced false-positivity that is encountered with other biomarkers, especially proteins, and is present in high quantities in plasma^32,34^. After being shed in circulation by apoptotic and necrotic tumor cells, ctDNA has a half life of about 16 mins to a few hours with a patient’s tumor burden determining the proportion of ctDNA found in serum. The usefulness of ctDNA analysis in patients with lung cancer and in patients with suspected lung cancer is broad and can be used for much more than a screening technique. ctDNA has been shown to be an effective tool for monitoring tumor burden and tumor response as well as providing a real time evaluation of metastatic disease^34^. In addition, ctDNA can be used to predict the prognosis based on the specific gene mutation and the effectiveness of certain chemotherapy^34^.

Because ctDNA has a sensitivity of 80%, a specificity of 96%, and is minimally invasive, our study found ctDNA could be used successfully as a screening technique throughout the population. As the biomarker is specific for genes only found in tumors, it can therefore reduce the chance of detecting a false-positive and provide a high likelihood of preventing a false negative. Further investigation into ctDNA would involve finding the specific segments of mutated genes in each form of lung cancer, and thus increase the sensitivity of ctDNA’s use as a biomarker. However, ctDNA should also be further researched for its other uses as well. Investigations into ctDNA’s predictive power of prognosis and the effectiveness of chemotherapies should thoroughly be researched as it has the potential to significantly decrease the morbidity and mortality of lung cancer.

Neovascular angiogenesis is a hallmark of cancer and abundant in neoplastic lesions. Thus, angiogenesis has an incredibly powerful potential to detect differentiation in a wide variety of cancers. Alfatide II is an upgraded version of the dimer RGD (Arg-Gly-Asp), specifically NOTA-E-(PEG_4_-c(RGDfk)_2_, which targets the integrin apha_v_-beta_3_; a neovascular maker^20^. However, the reciprocal remains true. In diseases such as TB there is a sharp decrease in the micro-vessel density, making Aflatide II a marker for both angiogenesis and vessel obliteration^20^. Thus, the ability of a ^68^Ga-Alfatide II CT/PET scan to identify such a wide-variety of diseases allows for an incredibly high diagnostic potential. However, the scan is not without limitations. As the scan can only detect angiogenesis, metastasis to the lymph nodes, bone and liver will have an inferior sensitivity to the ^18^F-FDG scan; this does not apply to brain metastasis as Alfatide II is able to penetrate the blood brain barrier^20^.

As the ^68^Ga-Alfatide scan has a sensitivity and specificity of around 85% and is minimally invasive, our study has found that this test would be appropriate for those at high risk for lung cancer or those suspected of lung cancer. However, it would not be recommended throughout the population as this biomarker screen requires a CT/PET scan with a contrast material. Thus, the test is expensive and counter-indicated in all forms of kidney injury. Further investigation should not be into Alfatide II itself, but rather what can be screened for in angiogenesis. While the CT/PET scan would have the greatest diagnostic spectrum of disease, a serum test would be ideal to lower cost and radiation exposure. Therefore, investigations into biomarkers for angiogenesis should be completed to find a biomarker synthesized only by tumor cells that can be found in serum, stimulating angiogenesis.

Paraneoplastic syndromes are a group of rare disorders triggered by an abnormal immune system response to cancerous tumors, referred to as neoplasms. This can be seen in some forms of lung cancers as well as other cancer types. The data obtained for this review did not contain research regarding biomarkers explaining this phenomenon. This is because it is only seen in late stages of cancer, while this review focused mainly on screening and prevention, which occur during developmental stages.

Additional future directions of this study would explore other journal platforms, such as google scholar or Scopus. This will allow a wider data size. In addition, the American Thoracic Society (ATS) journal was also a platform of interest that is yet to be explored for further findings on early screening methods for lung cancer. Other analytic points could also be used to further explore demographics by including smokers vs. non-smokers, and children vs. adults. Also, the use of immunotherapy treatments for lung cancers can be discussed.

Immunotherapy is currently a growing field that is scratching the surface of its potential. It is still being explored for oncology patients. The table below depicts current immunotherapies that have shown some forms of advancement in the case of lung cancer patients.^6^ An area of lung cancer immunotherapy treatment that requires further exploration involves comparing the efficacy between biomarkers successfully utilized in early detection and specific immunotherapy types within the same patient. This will better demonstrate response rates of immunotherapy in the treatment of Small Cell Lung Cancers (SCLCs) or Non-Small Cell Lung Cancers (NSCLCs).

Our study was limited by using articles that were only published within the United States and in the last 10 years, which constricted our sample size and the number of potential biomarkers that have been researched. We were also limited by the types of lung cancer each article chose to research as a biomarker could have been statistically significant in form of lung cancer that was not investigated. In addition, our study was limited by the possibility of false-positives and false-negatives in each of the articles we examined. Finally we are limited by any potential bias and limitations in each of the articles we examined.

## Conclusion

The study measured ctDNA and Ga-Alfatide II PET/CT scans as a means of screening for lung cancer with appropriate statistical values for measurement and data purposes. These are two biomarkers with highest sensitivity and specificity, thus indicating the chance of false-positive or false-negative are low. While these screening techniques cannot confirm the diagnosis of lung cancer, they have the potential to be indicated in any patient suspected of lung cancer or at high risk for lung cancer. Either of these biomarkers as screening techniques can increase early detection, thus decreasing mortality, provide indications into effective and/or ineffective treatments, and provide a clearer indication into the true morbidity of lung cancer.

Immunotherapy and chemotherapy are the most promising in comparison to other therapeutic options. By implementing these treatments, lung cancer surveillance before its development into advanced stages can be further improved. Although therapeutic effects can be increased with these standardizations, challenges may still arise due to constant changes during tumor growth. The complexity of various lung tumors could reduce the efficacy of these options due to therapeutic limitations, tumor resistance, or developed immune system tolerance to therapy. To overcome these challenges, it is important to develop screening methods for lung cancer, conduct further studies that analyze therapeutic response rates, and explore combination treatment if possible. Primary care physicians are vital in targeting challenges in treatments, as they can first assess the patient. Mortality rates of lung cancer have the potential of being lowered if the efficacy of immunotherapy and chemotherapy are increased. The improvement of screening methods, specifically the low-dose CT (LDCT) could also play an important role in decreasing death rates.

## Data Availability

All data produced in the present study are available upon reasonable request to the authors

## Acknowledgments

We would like to thank Dr. Arun Agnihotri, Sarah Moghari, Abrisham Akbariansaravi, and Shalom Aziague for their contributions to this paper.

## Notes

### Competing Interest Statement

The authors have declared no competing interest.

### Funding Statement

This study did not receive any funding

### Summary of Updates

The paper has been revised to remove the personal contact information of the authors

